# Bored, isolated and anxious: Experiences of prolonged hospitalization during high-risk pregnancy and preferences for improving care

**DOI:** 10.1101/2020.12.11.20247239

**Authors:** Adetola F. Louis-Jacques, Cheryl Vamos, Jessica Torres, Kathryn Dean, Emma Hume, Renice Obure, Ronee Wilson

## Abstract

Antepartum women who experience prolonged hospitalization are at greater risk for increased stress, anxiety, and depression. The purpose of this study was to examine women’s and healthcare staff’s experiences with prolonged hospitalizations during pregnancy and explore patient-centered intervention options. Guided by Social Support Theory, we conducted qualitative (focus group/interviews) and quantitative (surveys) methods. Patient survey respondents (N=57) were mostly white (77%), non-Hispanic (86%), and married (65%). Qualitative data (N=17) revealed challenges to women’s wellbeing related to: a) lack of knowledge; b) stress and anxiety; c) loss of independence; and d) social isolation in hospital; regarding concerns about hospitalization and competing demands at home. Suggested strategies for improving women’s experiences included interventions that involve family/friends, improved information sharing, and increased onsite services to facilitate a positive experience. This critical groundwork informed development of theory-based, patient-centered interventions to increase knowledge and enhance social support, among women who experience prolonged hospitalization during pregnancy.

## Introduction

Pregnant women with prolonged hospitalization are at greater risk for poor maternal mental health compared to their non-hospitalized counterparts as they must not only cope with the emotional stress of being pregnant but must also manage the composite stress of hospitalization. High risk categorization during pregnancy can also negatively affect the psychological health of antepartum and postpartum women and subsequent health and development of the infant (Garfield et al., 2015). High risk pregnancy is defined as a pregnancy that directly threatens the well-being or life of the mother or her fetus (NIH Office of Communications, 2017).

Pregnancy conditions defined as high risk can result in elevated neonatal morbidity and mortality, further increasing maternal stress (Howland, 2007). In addition, antepartum hospitalization has been associated with increased maternal psychosocial dysfunction (Correia & Linhares, 2007; Pichler-Stachl et al., 2016). The stress felt by hospitalized antepartum patients is multifactorial and composed of both modifiable and unmodifiable elements. Assisting hospitalized pregnant women in using coping strategies by increasing their knowledge and skills, and providing social support may help mitigate negative emotional impacts, ultimately reducing maternal stress and psychological dysfunction (Howland, 2007).

Previous studies on high risk pregnancies described the lived experience of hospitalized pregnant women using qualitative analysis (Denis et al., 2012; Leichtentritt et al., 2005; Rubarth et al., 2012), or using only quantitative analysis (Denis et al., 2012), did not explore potential intervention strategies (Pozzo et al., 2010), were mostly conducted outside the United States (Denis et al., 2012; Pozzo et al., 2010), and were not theory based (Denis et al., 2012; Leichtentritt et al., 2005; Pozzo et al., 2010; Rubarth et al., 2012). Thus, we developed a mixed methods study guided by Social Support Theory. Social support can encompass a variety of forms including informational support, emotional support, tangible support, and belonging support (Kent de Grey, 2018). Informational support includes the comfort that comes from knowledge or advice (Harvey & Alexander, 2012; Kent de Grey, 2018). Emotional support consists of the typical forms of support such as compassion, empathy, or caring (Kent de Grey, 2018). Tangible support is defined as the support that can be physically seen such as manual labor or financial aid (Kent de Grey, 2018). Lastly, belonging support takes into considering the support gained from a social network, such as a neighborhood, family, or friend group (Kent de Grey, 2018). The use of these social support constructs has been shown to improve health outcomes from cardiovascular disease to mental health stressors (Harvey & Alexander, 2012).

The overarching purpose of this mixed methods study was to explore patient and healthcare staff (1) experiences of prolonged antepartum hospitalization; and (2) opinions and preferences for proposed programming that will inform the development of patient-centered interventions. Using the mixed methods exploratory sequential design, the purpose of the quantitative methods was to help inform future interventions and refine the focus group and interview guide in the qualitative data collection (Canary, 2019). The purpose of the qualitative methods was to obtain detailed information on proposed interventions and understand the feasibility of implementing next steps.

## Materials and Methods

### Study Design

This was a mixed methods study utilizing exploratory sequential design. Mixed methods is a research method that utilizes both qualitative and quantitative data to investigate a hypothesis and formulate conclusions. The purpose of using mixed methods in the data collection, analysis, and interpretation phases of a study is to develop a more holistic view of the topic of interest (Shorten & Smith, 2017). Mixed methods research can be further broken down into sub-categories based on the way in which the qualitative or quantitative data drives the study. Exploratory Sequential design is one subcategory that uses quantitative data to drive targeted qualitative data collection (Cresswell, 2016). This methodology allows quantitative data to investigate an overarching idea, and the results from the quantitative data to guide qualitative data collection (Shorten & Smith, 2017).

### Sampling & Recruitment

***Pregnant/postpartum women*** were eligible to participate in a (1) survey and/or (2) focus group/interview if a) they were currently experiencing or had previously experienced a prolonged hospitalization during pregnancy (defined as 5 or more days); b) 18 years of age or older; and c) able to provide informed consent in English. Women were drawn from two sample populations. First, a local for purpose organization that provides support to women experiencing high risk pregnancy posted a link on their social media platform and website and sent an informational email to members. Second, research staff verbally introduced the study to a convenience sample of participants who were currently hospitalized. ***Key hospital staff members*** were eligible to participate if they held clinical, administrative, or other perinatal support roles to women experiencing a prolonged antepartum hospitalization at the affiliated teaching hospital. Staff members (e.g., physicians; nurses) were recruited through invitational emails. The study was IRB approved (#Pro00023050). Written or verbal informed consent was obtained from all individual participants included in the study.

### Data Collection

#### Quantitative Survey (Pregnant/Postpartum Women)

The survey instrument was available online or in-person (on paper) and was designed to determine if perinatal women were interested in participating in a support group, preferences for programming activities and support services that would be designed to provide them with the knowledge, skills and comfort to deal with a prolonged hospitalization during pregnancy. The survey solicited information on demographics, preferred group activities such as scrapbooking and pet therapy, and information on desired support services such as physical therapy and doula support. Before final administration, the survey was pilot-tested with the community partner to assess the wording, flow and clarity of the items. Results from the in-person and online surveys were utilized to guide focus group conversations and in-depth interviews.

#### Focus Group/Interviews (Pregnant/Postpartum Women)

A convenience sample of women who completed the survey were recruited for participation in a focus group and/or an in-depth interview. A focus group/interview guide was developed based on Social Support Theory, which posits that support from social networks and relationships offer a protective factor against susceptibility to stress (Glanz & Schwartz, 2008; Heaney & Israel, 2008). The focus group was subsequently refined based on quantitative survey results. Participants completed a brief demographic profile sheet including age, race/ethnicity, relationship status, pregnancy status, length of hospitalization stay, and reason for hospitalization. Women were asked about the quality and extent of *informational, emotional, instrumental*, and *appraisal* support that they experienced during their hospitalization. Suggestions were solicited on how to better foster these types of social support in a potential future intervention targeting pregnant women with a prolonged hospitalization. The focus group was conducted by a trained moderator; and the interviews were conducted by trained research team members. The focus group and interviews lasted approximately 90 minutes and 20 minutes, respectively.

#### Focus Group (Hospital Staff)

A focus group was also conducted among key hospital staff members to assist in providing a better understanding of the lived experiences of a prolonged hospitalization from an institutional and healthcare delivery perspective. This focus group also elicited feedback on the design and content of potential intervention elements, to ensure programming would be compatible with patient needs while being feasible and utilizing hospital infrastructure effectively. The staff focus group lasted approximately 60 minutes.

### Data Analysis

#### Quantitative

Frequencies and bivariate analyses (chi-square) were used to describe the responses and test for differences between survey groups (online vs. in-person survey administration) for the purpose of comparing the women who were still pregnant (in-person) and postpartum (online). Quantitative analysis was conducted using IBM SPSS 24.

#### Qualitative

All focus groups/interviews were audio-recorded and transcribed in preparation for qualitative data analysis (Canary, 2019). Additional interview notes were also taken by the moderator during the focus group sessions. Content analysis was performed on transcripts obtained from participant interviews and focus groups. Prior to coding the transcripts, the research team created a codebook containing a priori structural codes to identify and categorize themes based on Social Support Theory constructs. Transcribed focus group and interview data were coded by participant responses to each question. Two independent raters coded the transcripts in Microsoft® Word. Emergent codes were developed and incorporated as necessary throughout the data analysis process. The research team met to discuss emerging themes and choose illustrative quotes.

## Results

A total of 71 participants were enrolled for the mixed methods study, see Figure 1 for detailed information.

**Figure 1.**
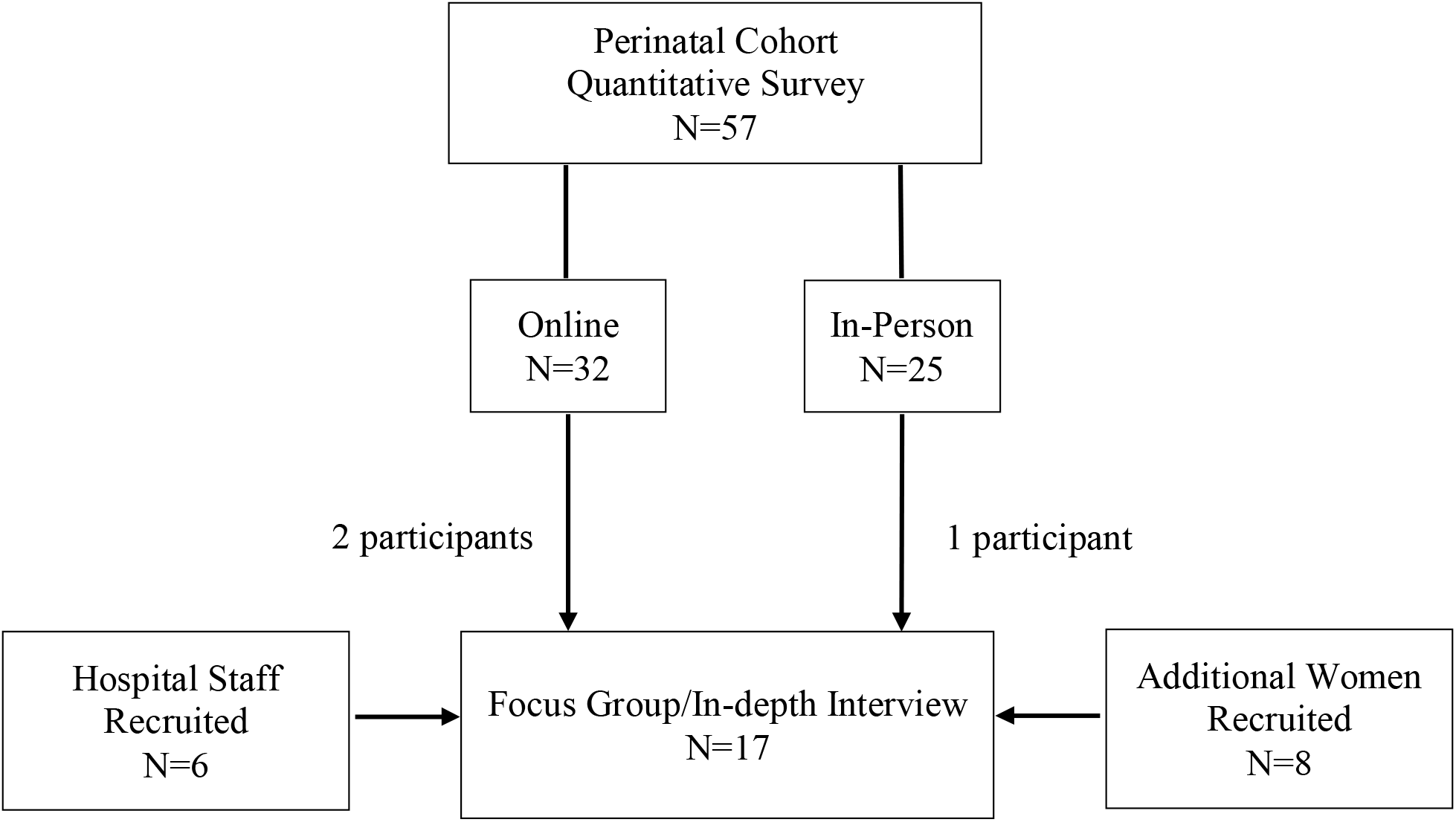
Quantitative & Qualitative Participants

### Quantitative Survey Findings *(Pregnant/Postpartum Women)*

In total, 57 women were surveyed either online (N=32) or in person (N=25) (Figure 1). The majority of women completing the survey in-person were still pregnant whereas the majority of women completing the survey online were postpartum. The mean maternal age was 31.3 years (SD ± 5.36). Survey respondents were primarily white (77.2%), non-Hispanic (86.0%), married (65.0%) and had at least some college credit (75.5%) (Table 1). Almost all participants (96.5%) were insured, with Medicaid accounting for 26.3% of coverage.

**Table 1:** Selected Socio-demographic Characteristics of Study Participants

| <u>Characteristic</u> | Total<br>(N=57) | In-Person<br>(N=25) | Online<br>(N=32) |  |
| --- | --- | --- | --- | --- |
|  | % | % | % | p-value |
| **Maternal age** |  |  |  | .008* |
| Mean (SD) | 31.30 (± 5.36) | 29.13 (±5.60) | 32.94 (±4.61) | |
| **Race** |  |  |  | .042* |
| Black | 8.8 | 16.0 | 3.1 |  |
| White | 77.2 | 60.0 | 77.2 |  |
| Other | 5.3 | 8.0 | 3.1 |  |
| No Response | 8.8 | 16.0 | 3.1 |  |
| **Ethnicity** |  |  |  | .037* |
| Hispanic | 14.0 | 24.0 | 6.3 |  |
| **Education** |  |  |  | .010* |
| <High School | 5.3 | 8.0 | 3.1 |  |
| HS/GED | 14.0 | 28.0 | 3.1 |  |
| Technical | 5.3 | 8.0 | 3.1 |  |
| Some College | 15.8 | 20.0 | 12.5 |  |
| Associate Degree | 8.8 | 12.0 | 6.3 |  |
| Bachelor Degree | 28.1 | 16.0 | 37.5 |  |
| Graduate Degree | 22.8 | 8.0 | 34.4 |  |
| **Marital Status** |  |  |  | .001 |
| Married | 65.0 | 44.0 | 87.5 |  |
| **Insurance** |  |  |  | .689 |
| Yes | 96.5 | 96.0 | 96.5 |  |

*\*values significant at $\alpha=0.05$*

Attitudes and perceptions on potential programming and services are presented in Table 2. There were significant differences between participants who answered the online survey and those who completed the paper survey in person. Women who completed the survey online were slightly older and more likely to be white, non-Hispanic, college educated and married when compared to women who completed the paper survey.

**Table 2.**
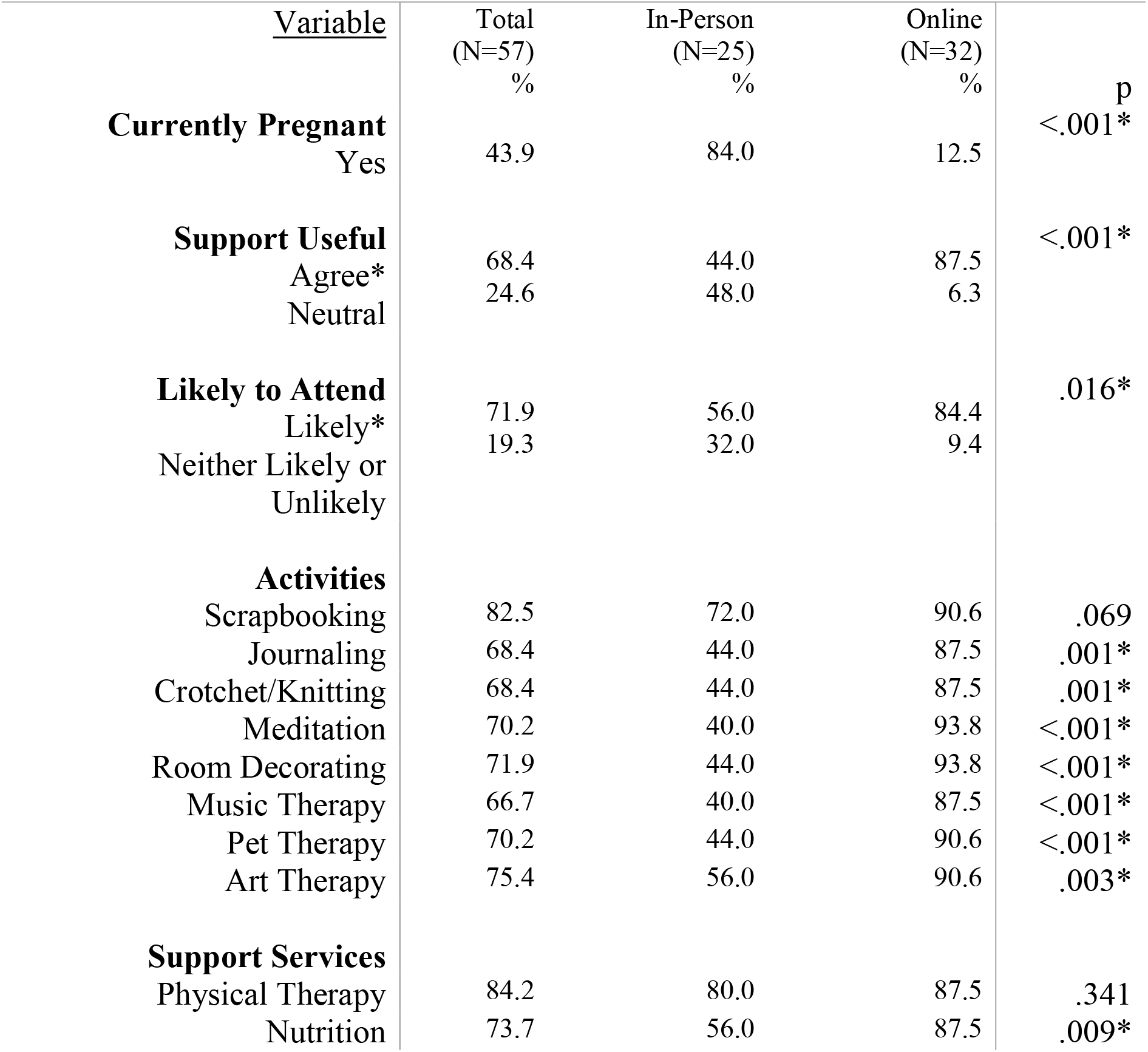
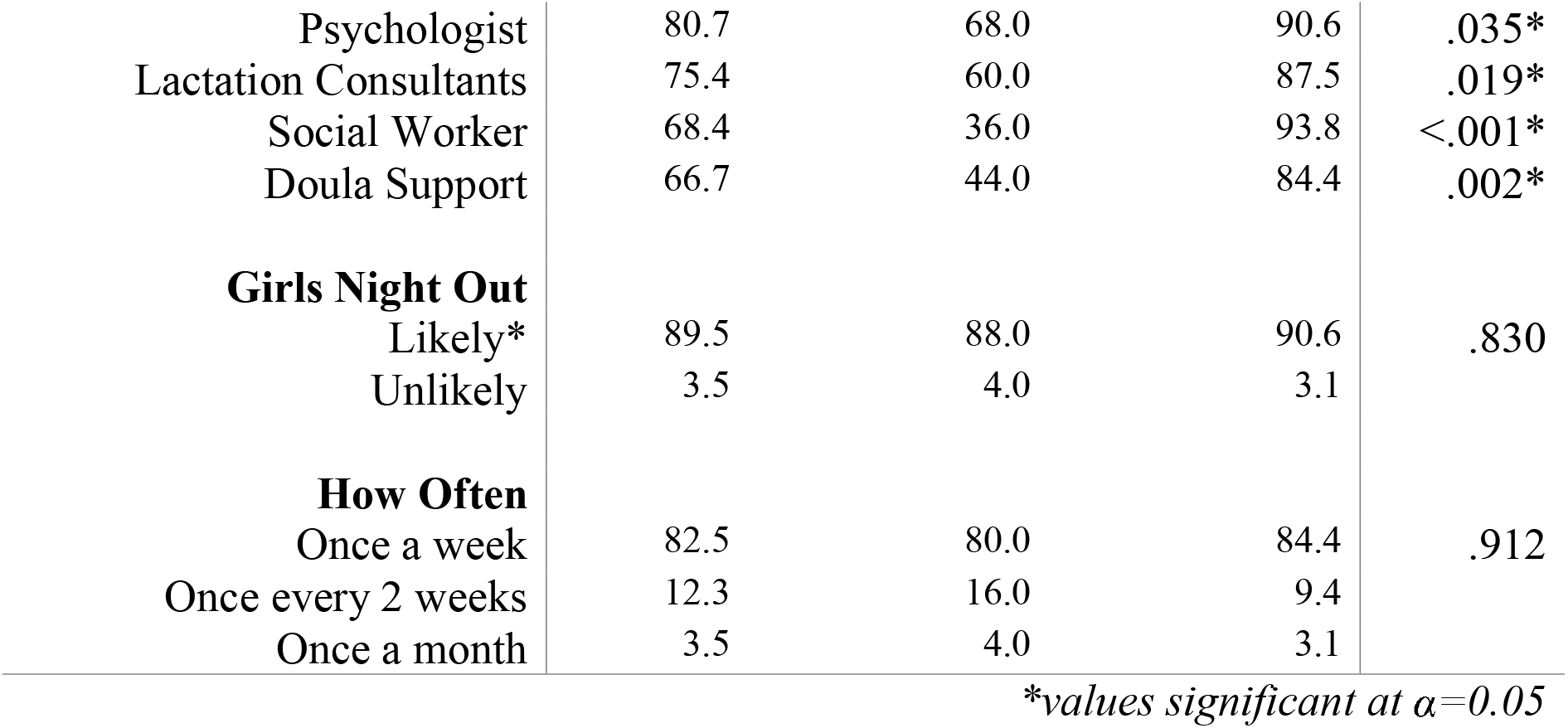
Attitudes and Perceptions on Support Groups and Potential Coping Activities

Preferences regarding participation in group activities and support services also differed by survey mode of delivery. Women who completed the survey online overall showed an interest in all of the suggested activities and support services. On the other hand, responses from the paper surveys indicated minimal interest in suggested activities and support services except scrapbooking, physical therapy, and psychology services.

#### Focus Group/Interviews Findings (Pregnant/Postpartum Women)

Eleven women participated in the qualitative component (Focus group/in depth interviews) (Fig. 1). The mean age of the women who participated in the qualitative component was 20.00 ± 6.10 years with a range of 18 to 40. The racial/ethnic distribution was 54.5% white, 36.4% black, 9.1% American Indian/Alaska Native and 100% non-Hispanic. The majority of participants were married or living with their partner (72.7%) and 63.6% were pregnant during the interview. All participants had at least a high school education or GED and approximately 81% has enrolled in college or obtained a college degree. Based on the qualitative data, several themes emerged including: fear of the unknown; loss of independence; boredom; poor quality and limited food options. These experiences are described further below.

#### Lack of knowledge and poor communication

Participants described the lack of knowledge that they had regarding their medical condition(s), and the associated medical procedures, treatment plans and potential complications. One woman explained: “*So I knew about breastfeeding, but not the steroid shots, kind of did research, but why are they doing different things?*” Participants also indicated they received too much print information which was not necessarily helpful. One woman suggested that staff should “*go through it, like the highlights of this, so the person knows this is helpful, this is not helpful, what’s in it. After I sat there. I was like why did I read all of this*.”

In addition, participants described not understanding the medical terminology that the providers used during their discussions.

> *Sometimes I feel like the doctors didn’t explain everything correctly what was going on and you felt lost and there’s some people that don’t understand medical knowledge*.

Participants also described not knowing what to expect with regards to the labor and delivery process and postpartum periods.

Moreover, participants did not appreciate being told what to do without an explanation or without being involved in the decision-making process. Communication barriers meant that at times when the patients did not understand what the doctors said, they resorted to finding out information from other sources which may be incorrect. For example, as one participant stated: “*I know I like to read on what is going to happen and I go to Google sometimes which is very bad*”. Compounding these information deficits and the emotional impacts were participants’ inability to explain to their family members what was happening to them.

#### Stress and anxiety

Participants also described high levels of anxiety related to not knowing what to expect concerning their pregnancy and delivery and health of their baby. This was commonly described by participants as “*the unknown*.” A cascade of events and emotions, where everything is okay one minute and then complications arise, leading them to get rushed to a different room for intervention, further contributed to high stress levels. At the core of these experiences was the constant worry about when they might go into labor and whether their baby would be healthy.

> *You never know if your baby is going to come early or not. You are never prepared for that. I never know what’s going on before they come rushing in*.
>
> *It was the unknown, how was he going to come out, how small or big would he be*.

#### Loss of Independence

Participants reported feeling a loss of independence and autonomy. They explained how they had to contend with hospital rules restricting their movements and activities. This loss of freedom was isolating, frustrating and felt like “*jail time*” as one patient noted. The confinement and being told what do negatively impacted their sense of control and independence. These constraints and limitations not only pertained to their immediate hospital environment but also to decisions within their household. They expressed feelings that their “*life was on hold*.”

> *I’m a big control person and a control freak and so not being able to control what’s going on what’s in my house when I’m in the hospital it is very kind of overwhelming and king of upsetting really*.

#### Isolation and Boredom

Isolation was a major emotional impact that women experienced during their antepartum hospitalization. Participants discussed being isolated from people in general, from other pregnant women, and from their families. Some families did not live close by or were busy with the day-to-day operations and functioning of running and family, and participants greatly missed being able to see their loved ones.

> *It really sucks. I feel like I don’t enjoy my pregnancy as much because I lose out on like a lot of different things I did you know beforehand. I’m not around my family. That sucks. And it’s just hard, being alone is really hard*.
>
> *The hard part is, is being away from, you know, your family. For me, I have a child at home so it’s really hard for me to think about ok who is going to watch her. How is she eating? How is she doing? Is she you know that mommy still loves her and mama’s here? And having to worry about your new baby and make sure your new baby is ok*.

Participants also explained how not knowing other patients going through similar experience, doing the same thing every day, and hospital restrictions on movement and activities led to boredom. As one of the patients aptly described the situation, “*There are only so many movies to watch and so much TV and you get really bored and then it makes you think more and you get more depressed. It’s hard to be in the same hospital room forever and ever*”.

#### Quality of Stay

Participants described elements in their social and physical environment that either helped them cope with their prolonged stay or negatively impacted their experiences. Engaging in relaxation techniques, such as meditation and listening to music, were identified as being useful in helping them cope with their time at the hospital. Forms of social support from friends and family were also described as mitigating the stress from their situation. Often these were forms of instrumental support where their family brought in resources to aid in their comfort.

> *My husband’s stepmom brought a lamp, didn’t have to have fluorescent light all the time, own pillows, not much space to set things, had my computer, kindle, phone, there for a while, own snacks, had a little fridge family brought, things that make me comfortable, brought outside food, people would bring stuff*.

Given their extensive confinement in the hospital, food availability and quality were also mentioned as negatively impacting their stay. Participants found the menu options offered through the hospital to be limited.

#### Life Doesn’t Stop

The hospital stay created financial and logistic issues, and participants were left to solve problems external to their medical condition(s), such as how the bills would be paid and how they were going to take care of other children who were back home. These life demands were further escalated among participants who had limited financial and social resources. With at least one participant, there were concerns regarding whether she would still have her job.

### Focus Group Healthcare Staff Findings

Six hospital providers/administrators with varied experience and expertise including one maternal-fetal medicine physician, one social worker and four nurses, participated in the staff focus group. Two of the nurses were managers, and two were lactation consultants. All participants were female, white, non-Hispanic. The age ranged from was 18-59 with 5 out of 6 providers having more than 10 years of experience in the healthcare field.

The providers in this study described their perceptions of the experiences of high-risk pregnant hospitalized women, as well as their own experiences when working with this population. Three themes were identified during the staff focus group: (1) balancing home life vs. hospital life (patients); (2) work burden (staff); and (3) preferences and feasibility of future programming.

#### Balancing home life vs. hospital life

Hospital staff members witnessed how difficult it was for antepartum hospitalized women to carry out home responsibilities such as childcare and financial duties. For example, one staff participant said, “*I remember having to help a woman translate in Spanish, I told her you have to stay here, you have to be admitted, she says, I can’t stay here, I have all these things, didn’t understand I was being admitted, helping her coordinate everything, especially women who don’t have many resources, someone they are comfortable with taking care of child*.”

#### Work Burden

Participants explained how they and other staff members individually perform both high-risk rounds and assess the types of support services their antepartum patient may need, but only if they had extra time available. One participant said, “*I’ve had time to do it, but if I’m busy then maybe I can’t at that time*.”

#### Preferences and Feasibility of Future Programming

One provider even suggested having an antepartum coordinator to help orient antepartum patients during clinical flow. She said “*would like a coordinator, who orients them, go over menu, what would help, reach out to social worker to help pay bills, need to breastfeed baby when born, let me get the lactation, public transportation, feeling stressed, how about music, or pet*.”

### Suggestions for proposed intervention based on Social Support Theory (All Participants)

Overall, participants identified the desire to have (1) informational and (2) social programming (see Table 3). Informational programming included providing information in a clear, user-friendly manner about medical procedures, including tests that were routine and advanced given their high-risk situation. Information with regards to general infant care, similar to what is discussed during prenatal classes, was also suggested. In addition, both patients and hospital staff members believed that information specific to their prolonged hospitalization should be condensed and explained carefully. Language and cultural differences should be considered.

**Table 3.** Proposed Intervention from Participants

| **Table 3. Proposed Intervention from Participants** |  |
| --- | --- |
| **Theoretical Constructs** | **Input** |
| **Informational/Appraisal Support** | “folder with tabs, what to expect for NICU, tab for breastfeeding, tab for surviving hospital stay, tab for HRH, tab for healthy start, tab for social services...more condensed... opposed to different people coming in” |
| **Instrumental Support** | “I think there should be more options for food because once you eat a cheeseburger for 9 days,....., it kinda gets really boring and the best part of my whole stay was when my mother took me downstairs and I got to pick what I wanted to eat” |
| **Emotional Support** | “I definitely think it would be nice to communicate with the different women on the floor you know like a support group or like a once a day meeting to share their experience. I know that tends to help a lot”<br>“[I] wish there was more time for activities, get exposure to other people; I’m in a room, door shut most of the time, not seeing another patient.”<br>“You guys take away the freedom that I have. I’m strapped to the bed. I can’t get up I can’t shower. You barely let me |
|  | eat a couple of days” |

Antepartum women expressed interest in having social gatherings with other women experiencing similar situations at the hospital. Suggested ideas for social programs included arts and crafts, manicures, pedicures and prenatal massage. For these types of activities, women explained that they preferred to mingle with other women while doing activities without the involvement of family members. It was recommended that various activities should be offered to appeal to a broad array of interests and needs. Lastly, participants stressed that programs should be scheduled during a time that does not conflict with the routine clinical flow as meeting with their providers and hearing any updates on their case was the most important priority to them.

### Preliminary Interventions Developed

Using this formative research, two interventions were developed based on the results from the quantitative and qualitative feedback from participants. Input from both patients and providers were taken into account when designing (1) Informational Programming and (2) Social Programming.

### Informational Programming

The Informational Programming intervention created was an *Informational booklet* to be given to patients during their antepartum stay at the hospital. The purpose of this booklet is to reduce anxiety and uncertainty by providing information on anticipated aspects of their hospital stay such as: medical terminology explanations, medical procedure explanations, medical team introductions, childcare, and food delivery options. This resource is available in English and Spanish.

The *Informational booklet* has been reviewed by patients, maternal fetal medicine attending physicians and fellows, general obstetricians, residents, labor and delivery nurses, antepartum nurses, nurse managers, social work, physical therapy, the *patient experience* and *marketing* divisions of Tampa General Hospital. The booklet contains the following information:

1. Introduction to the hospital-units where our patients may stay, nursing team, types of monitoring (maternal and fetal), reasons for transfer between units
2. Teams involved in their care
3. Expectations during admission are addressed including; initial intake in triage, fetal monitoring (intermittent versus continuous), continuity of prenatal care during admission if indicated, neonatal ICU team consultation, and timing of daily rounds
4. Common conditions and reasons for admission
5. Common medications administered during a high risk pregnancy
6. Amenities discussed include dining options in the hospital as well as nearby restaurants and grocery stores, internet, lodging for family and *Ronald McDonald House*, options for childcare, chapel, gift shop
7. Physical therapy handout developed by physical therapists at the hospital with pictures and descriptions of exercises participants can do while in their hospital room

Preliminarily participants have found the booklet to be beneficial. Further formal evaluation is ongoing to assess the impact the *Informational Booklet* has on the mental well-being of antepartum patients.

### Social Programming

The Social Programming intervention was the development of a support group for antepartum patients. An interdisciplinary team consisting of lactation consultants, physical therapists, recreational therapists, integrative medicine, social workers employed by the hospital was assembled. In addition to these hospital employees, High Risk Hope, a for-purpose organization for women and families experiencing high risk pregnancies was also included. Each week, a different group hosted a one-hour meeting. During the first half of each meeting, there was an informational session about hospital resources, a learning topic, or activity and then the second half was an informal opportunity for the women to socialize with one another. The nursing staff helped advertise and escort patients to and from the sessions.

## Discussion

Antepartum women for whom prolonged hospitalization is prescribed, experience increased stress and anxiety. Themes emerging from our study such as isolation, home-hospital life balance, unwanted dependence and increased anxiety are similar to previous findings from Rubarth and colleagues (Rubarth et al., 2012). In addition, our findings regarding poor and inefficient communication in some circumstances between the patients and providers have also been reported (Pozzo et al., 2010). Patients and their families felt overwhelmed with a lot of information received, and it was ineffective at meeting both informational and emotional needs. Moreover, healthcare staff desired to do more but did not always have enough time to spend with patients to address complex social and emotional needs. Additionally, healthcare staff in our study highlighted the need for an antepartum coordinator to structure and coordinate existing resources available at the hospital, or create new patient-centered activities.

Our findings suggest that the desired intervention should be informational as well as interactive without disrupting clinical flow. Patients desired to be informed and involved in the medical decision-making process with simple, thematically structured and well explained information. Furthermore, participants desired an intervention to include social sessions where women can meet to provide peer-to-peer support and encouragement; as well as inclusion of therapeutic activities such as art therapy and scrapbooking. With this in mind, this formative study informed the design and development of the Informational Programming and Social Programming interventions.

These findings must be considered in light of noted limitations. First, the sample size for the qualitative methods was small and thus it is unclear whether data redundancy or saturation was met. Second, the sample was not equally stratified between those who were currently hospitalized or were previously hospitalized at time of data collection. The differences between the participants who completed the survey online and those who completed in person could be attributed to the fact that the majority of online participants were postpartum whereas the majority of in-person participants were pregnant. The responses indicated a more enthusiastic response in retrospect from women whose high-risk pregnancy was resolved. The differences also reveal some apathy or slight disinterest in the support sessions from women currently hospitalized. However, if effectively implemented women would participate in the program. Lastly, participants received antepartum/postpartum care in a large metropolitan city where there are resources and medical expertise available, such as maternal-fetal medicine physicians and level III neonatal intensive care. Thus, findings cannot be generalized to other regions or other women who have experienced a prolonged hospitalization during pregnancy.

Conversely, this study posits several strengths. This was a mixed method study, which was able to quantitatively describe thoughts and preferences among participants, while elucidating rich contexts to provide a deeper understanding of participants’ experiences. Furthermore, individual interview and focus group data revealed reoccurring themes. Moreover, both providers and patients’ perspectives were utilized and triangulated to inform the development of patient-centered interventions within the confines of antepartum hospitalization.

## Conclusions

There is need for structured interventions for hospitalized women in the antepartum period. When pregnant women are hospitalized, appropriately the medical needs are priority. However, attention should also be directed to the complex social and emotional needs of pregnant women experiencing a prolonged hospitalization. Hospital should ensure the care for pregnant women experiencing a prolonged hospitalization is patient centered and holistic. There is limited research on interventions to support this population. This formative work is an example of how to engage women in the development of interventions that support them and their families during a difficult period of their lives.

## Data Availability

The data referred to in this manuscript is available upon request.

